# Developing RT-LAMP Assays for Detection of SARS-CoV-2 in Saliva

**DOI:** 10.1101/2021.04.25.21256085

**Authors:** Xin Huang, Gongyu Tang, Nahed Ismail, Xiaowei Wang

**Author notes:** Corresponding author: Xiaowei Wang.

## Abstract

The coronavirus disease 2019 (COVID-19) caused by SARS-CoV-2 has killed millions of people worldwide. The current crisis has created an unprecedented demand for rapid test of SARS-CoV-2 infection. Reverse transcription loop-mediated isothermal amplification (RT-LAMP) is a fast and convenient method to amplify and identify the transcripts of a targeted pathogen. However, the sensitivity and specificity of RT-LAMP were generally regarded as inferior when compared with the gold standard RT-qPCR. To address this issue, we combined bioinformatic and experimental analyses to improve the assay performance for COVID-19 diagnosis. First, we developed an improved algorithm to design LAMP primers targeting the nucleocapsid (N), membrane (M), and spike (S) genes of SARS-CoV-2. Next, we rigorously validated these new assays for their efficacy and specificity. Further, we demonstrated that multiplexed RT-LAMP assays could directly detect as low as a few copies of SARS-CoV-2 RNA in saliva, without the need of RNA isolation. Importantly, further testing using saliva samples from COVID-19 patients indicated that the new RT-LAMP assays were in total agreement in sensitivity and specificity with standard RT-qPCR. In summary, our new LAMP primer design algorithm along with the validated assays provide a fast and reliable method for the diagnosis of COVID-19 cases.

## Introduction

The pandemic of COVID-19 has killed over 2.7 million people worldwide, including over 545,070 in the USA as of March 25, 2021. More than 124 million people have been infected worldwide. Each day, hundreds of thousands are added to the numbers (while this manuscript was being prepared). Rapid testing to identify the SARS-CoV-2 positive population followed by quarantine is important to curb the pandemic. Thus, the current crisis has created an unprecedented demand for rapid tests with high sensitivity and specificity for point-of-care diagnosis. Vaccine distribution has curbed the pandemic significantly, although it is also expected that the virus will be present in communities in the long term ^1^.

To detect SARS-CoV-2, many research groups have developed new diagnostic assays based on various technology platforms. Among them, nucleic acid detection by reverse transcription quantitative PCR (RT-qPCR) is considered the gold standard for virus diagnosis. In addition, there are other alternative nucleic acid detection methods developed in recent years, providing fast diagnostic results ^2^. However, these newly emerged methods generally have inferior diagnostic performance compared with RT-qPCR ^3^. Among these methods, loop-mediated isothermal amplification (LAMP) was developed by Notomi T et al. in 2000 ^4^. Since then, LAMP has been widely adopted for the detection of many pathogens, such as malaria ^5^, salmonella ^6^, influenza virus ^7^, dengue virus ^8^, Chikungunya virus ^9^, and Zika virus ^10^. Although multiple groups have attempted to apply the LAMP technique to SARS-CoV-2 detection ^11-13^, there are still major unresolved technical challenges preventing its application in clinical settings.

The advantage of LAMP assays mainly lies in their fast turnaround time and simplicity in assay setup. Specifically, LAMP does not require high-end instruments, making it widely accessible worldwide including developing countries. Moreover, LAMP works well even for a variety of unpurified samples, which is important for convenient and fast assay setup. Another major advantage is that LAMP readout can be colorimetric or turbidity changes, easily visualized and recorded by a phone camera. In addition, being performed under lower temperatures than PCR, LAMP can be easily combined with the RT reaction (RT-LAMP), directly detecting target RNA without a separate RT step. In this way, the total reaction time can be greatly shortened. Therefore, LAMP is a powerful tool for point-of-care diagnosis with wide applications ^2^. Despite these advantages, current LAMP assays are regarded as inferior to RT-qPCR in detection sensitivity and specificity ^11, 14^.

Compared with PCR, LAMP is still an emerging technology with major unresolved issues. It is a major challenge to design robust LAMP assays for pathogen detection. A typical LAMP assay requires six primers targeting eight regions of the target sequence (Fig. 1). At present, few tools are available publicly for LAMP primer design, and available tools did not sufficiently consider the complexity of LAMP reactions. For example, one major issue is related to high level of unintended primer cross-reactivity, given that a set of six primers are included in a single LAMP reaction. This frequently leads to false-positive results as reported by many investigators ^11, 15-18^.

**Figure 1.**
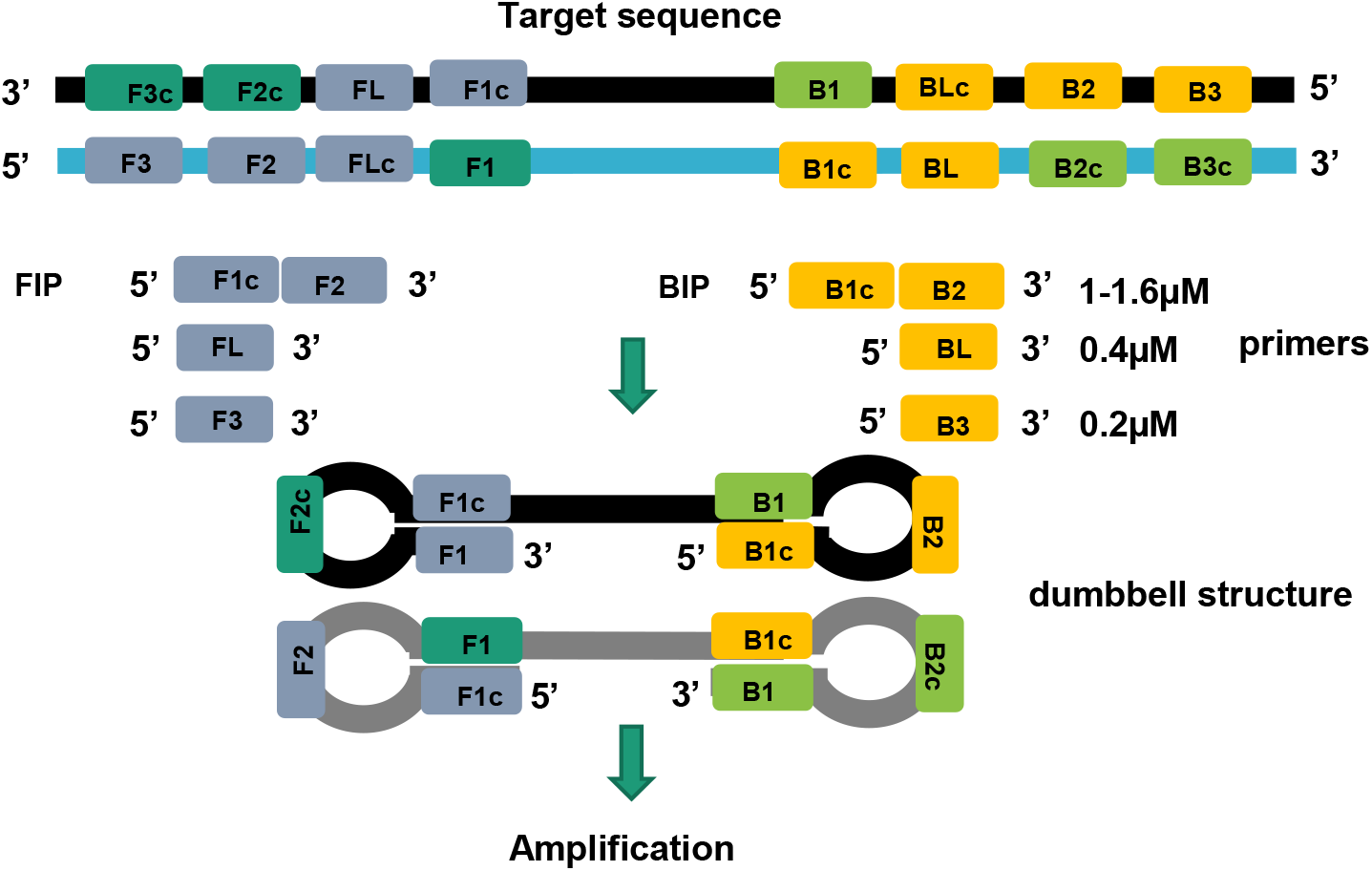
The LAMP assay contains six primers annealing to eight regions of a target sequence.

In this study, we developed an improved bioinformatics algorithm to design LAMP primer sets targeting the nucleocapsid (N), membrane (M), and spike (S) genes of SARS-CoV-2. The performance of these primer sets was rigorously validated experimentally. A combination of the best primer sets could reliably detect as low as a few copies of SARS-CoV-2 RNA in unpurified specimens (i.e., saliva) without the need of RNA isolation. This represents significant improvement over other reported LAMP assays ^11-13^. Finally, further testing using simulated saliva samples as well as patient saliva samples indicated that our new assays were as effective as standard RT-qPCR in SARS-CoV-2 detection. Importantly, our assays have unique advantages over RT-qPCR due to their fast and direct detection of viral RNA in saliva with colorimetric readout, making it ideal for inexpensive point-of-care diagnosis. Our new LAMP design method as well as experimentally validated assays provide a valuable resource to fight the pandemic of COVID-19.

## Results

### Optimization of the primer length to improve LAMP efficiency

One major goal of our study was to improve the efficacy of LAMP assays for SARS-CoV-2 detection. During the LAMP reaction, the amplification stage is initiated with the formation of a dumbbell structure, in which F1/F1c (or B1/B1c) and F2 (or B2) form the stem and loop, respectively (Fig. 1). As the first step, two long inner primers, FIP (F1c+F2) and BIP (B1c+B2) are annealed to the target to initiate the LAMP reaction. In general, long oligo primers have relatively poor annealing efficiency due to the potential formation of secondary structures. The poor annealing efficiency of long oligo primers has been extensively documented in numerous PCR studies ^19^. Thus, we reasoned that the efficacy of LAMP assays could be improved by shortening the FIP and BIP primers. To this end, we determined the optimal length of the stem region (F1c or B1c) within the primers. As shown in Table 1, we first designed N1(−3), and N2(−5) assays, which had shortened stems (as compared to standard design) to target the N gene of SARS-CoV-2. The shortening of the stems resulted in significantly improved TT values in comparison to that of original N1 and N2 assays (Fig. 2A). Thus, it was clear that standard LAMP design guidelines for the dumbbell structure, as recommended by previous studies ^4, 20^, should be further optimized.

**Table 1.**
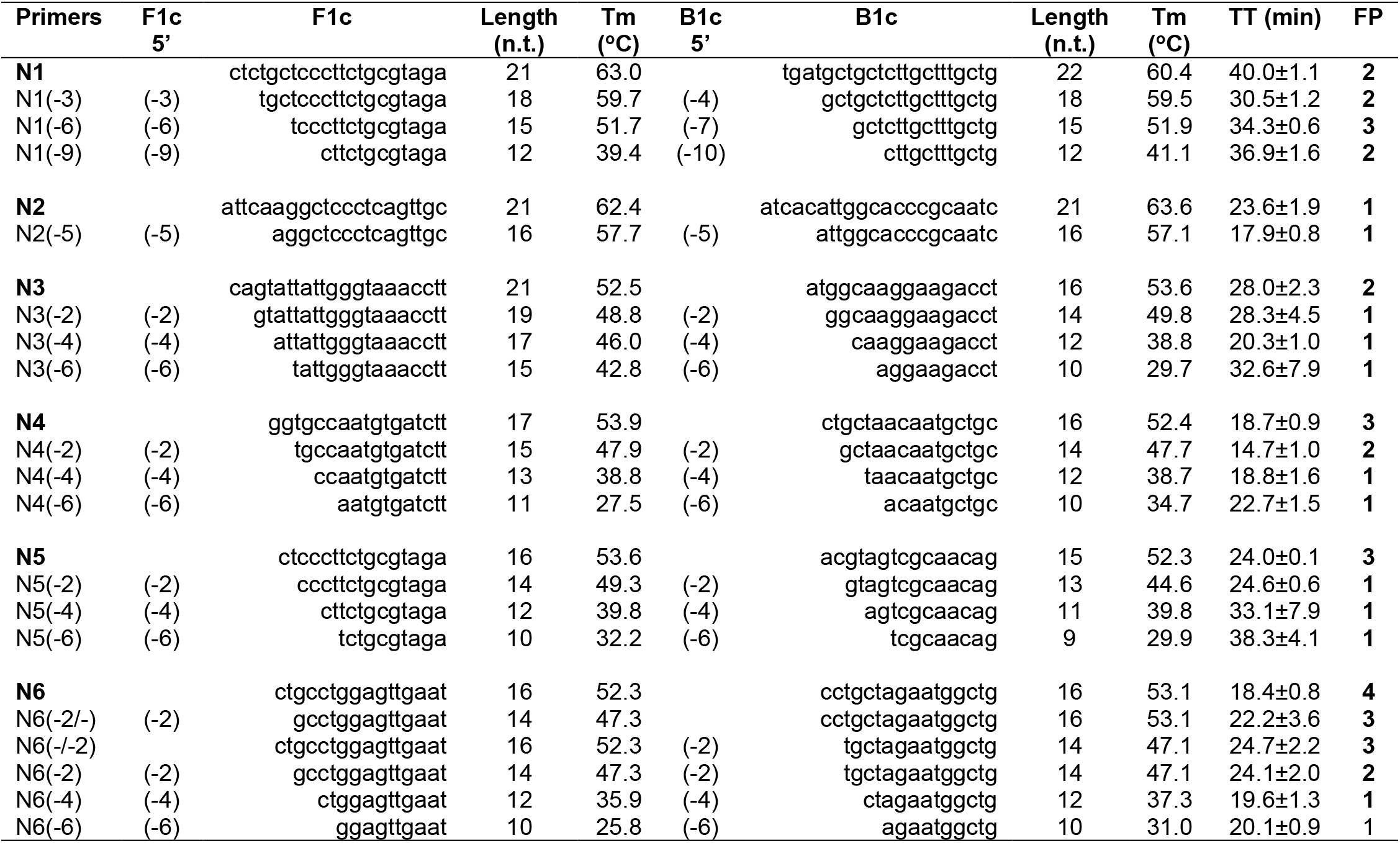
Effects of shortening the loop stem (F1c and B1c) on amplification efficiency and specificity.

**Figure 2.**
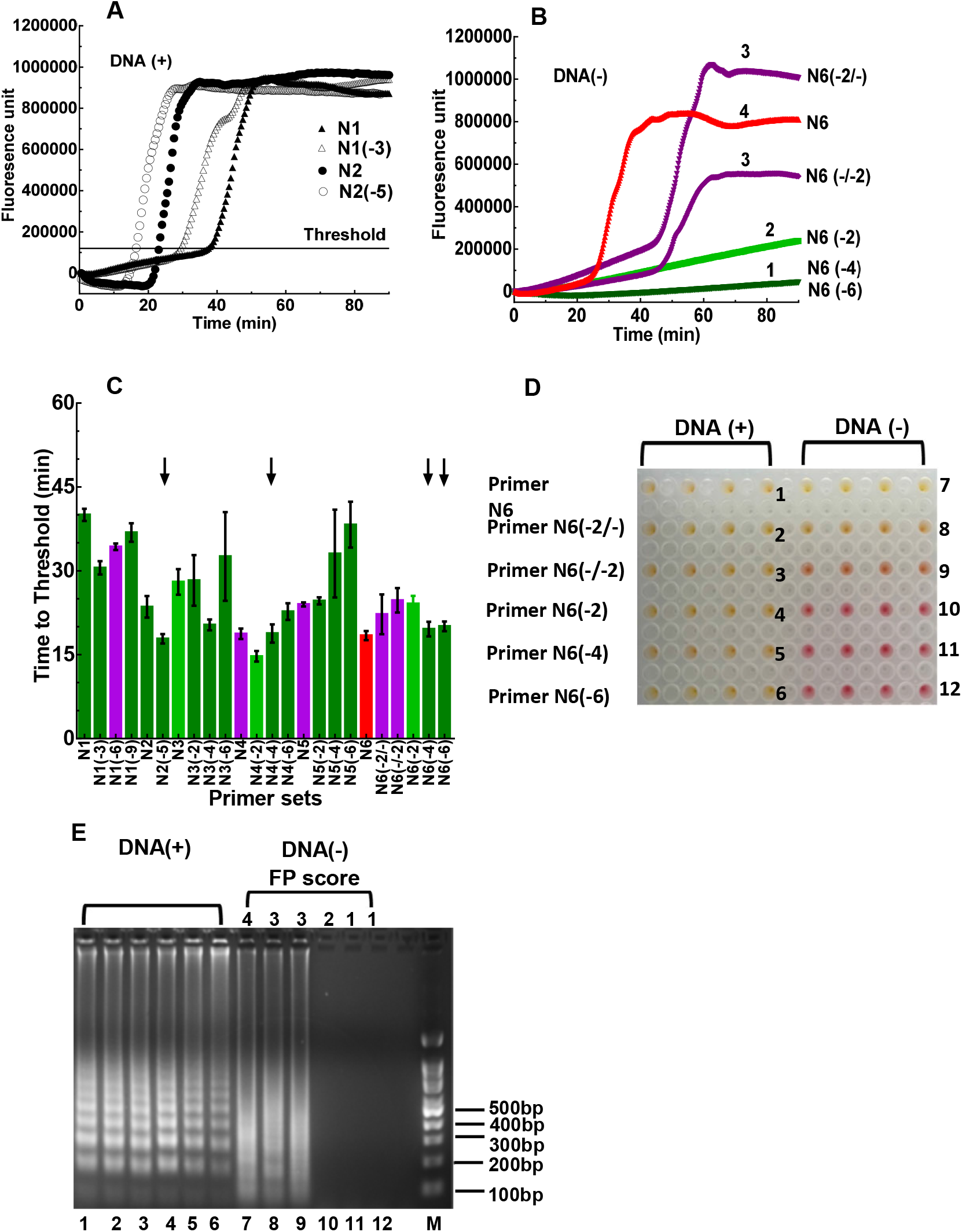
Assessing the sensitivity and specificity of the LAMP assays. DNA plasmid containing the N gene of SARS-CoV-2 was used as template for LAMP amplification. **(A) Amplification plots of LAMP reactions**. Each curve represents an average of 3 independent measurements. **(B) False-positive signals produced by the N6 and N6-derived assays**. The LAMP assays were performed in the absence of the N gene template. The false-positive (FP) signals were evaluated using the FP score (score range 1-4). 1) Dark green: no or minimal fluorescence signals at endpoint; 2) light green: low but detectable signals at endpoint; 3) purple: signals detected during late stage of the reaction (45-90 min); 4) red: signals detected during early stage of the reaction (< 45 min). (**C) The TT values and FP scores of the assays targeting the N gene**. The same color scheme as in Fig. 2B was used to assess false-positive signals as determined from no-template control reactions. Primer sets N1 to N6 as well as their derived assays, which contained identical F2, B2, FL, BL, F3, and B3 primers but shortened F1c/B1c primers. The arrows indicated selected assays with best overall performance in LAMP efficacy and specificity. (**D) Colorimetric changes of the reactions using the N6 and N6-derived primer sets**. Left panel: with DNA template included; right panel: no DNA template. (**E) Gel electrophoresis analysis of the LAMP products**. The same LAMP products as described in Fig. 2D were analyzed on DNA agarose gel. On-target products showed typical DNA ladders (lanes 1-6) while non-specific products with high FP scores showed DNA smears (lanes 7-9).

To comprehensively determine the optimal length of the stem region, we further designed twenty primer sets with variable F1c and B1c lengths (derived from the N1 to N6 assays). The testing results and the sequences of these primer sets are summarized in Table 1 and Supplementary Table S1, respectively. Both the length and melting temperature (Tm) of the stem region were important determinants of assay efficiency. While stem shortening helped improve TT values as compared to the standard design, excessive further shortening of the stem region was detrimental as a result of significantly lowered Tm value. Based on these 24 LAMP primer sets, observed optimal stem length was in the range of 12-17 bp with Tm >45°C.

### Assessment of non-specific LAMP products

Besides amplification efficiency (as represented by TT), another major consideration of LAMP performance is the specificity of the assays. False-positive products are commonly observed in LAMP reactions even in the absence of nucleic acid targets, as reported by numerous studies ^11, 15-18^. For example, multiple LAMP assays derived from the N6 assay (Table 1) showed various levels of false-positive readout in the absence of the target template (Fig. 2B). Thus, it is crucial to design primer sets with low cross-reactivity to avoid false-positive amplification. To this end, we categorized potential false-positive (FP) reactions into four levels (FP scores 1-4) during primer screening with representative examples listed in Fig. 2B: **1)** FP score of 1, no or minimal fluorescence signals at the endpoint, as represented by N6(−4) and N6(−6) assays; **2)** FP score of 2, low but detectable fluorescence signal at the endpoint, as represented by N6(−2); **3)** FP score of 3, high fluorescence signals detected during the late stage of the reaction (45-90 min), as represented by N6(−2/-) and N6(-/-2); **4)** FP score of 4, false amplification observed during the early stage of the reaction (<45 min), as represented by N6. By employing this color-coded scoring scheme, we were able to concurrently evaluate both the efficiency and specificity of the LAMP assays (Fig. 2C).

LAMP assays with various FP scores had distinct colorimetric readouts and electrophoresis patterns in the absence of target DNA (Fig. 2D&E). Specifically, for assays with an FP score of 1 or 2, no colorimetric change was observed (Fig. 2D, pink wells in the right section for N6(−2), N6(−4), and N6(−6) assays). In addition, no amplification product was observed by gel electrophoresis (Fig. 2E, lanes 10-12). For assays with a score of 3 or 4, non-specific products were observed by both colorimetric changes [Fig. 2D, yellow or orange wells in the right section for N6, N6 (−2/-), and N6(-/-2)] and gel electrophoresis (Fig. 2E, lanes 7-9). Interestingly, the size distribution of non-specific products was distinctively different from that of on-target products (Fig. 2E lanes 1-6 vs. 7-9). To maximize assay specificity, in our study, only primer sets with an FP score of 1 were further evaluated with the goal of developing robust assays for SARS-CoV-2 diagnosis.

To understand the mechanism underlying the false-positive reactions, we further analyzed non-specific products from the N6 assay. The N6 primer set was prone to false-positive reaction, having an FP score of 4. Further analysis of the non-specific products by electrophoresis showed that false-positive reaction happened in an FIP/BIP dependent manner (Supplementary Fig. S1A). The false-positive reactions could occur even in the absence of FL, BL, F3, and B3 primers (lanes 5-8, FP 3-4), but not in the absence of FIP/BIP primers (lanes 9-12). Thus, it was clear that the FIP/BIP primers in the N6 assay were responsible for the false-positive products. We further characterized the false-positive products by high-throughput sequencing. To this end, PCR primers with F2 or B2 sequence at the 3’-end were used to amplify non-specific LAMP products from the N6 assay (Supplementary Table S2). Sequencing showed that both FIP and BIP primers were incorporated into the non-specific products, with new linker sequences (1-5 bp) inserted between FIP and BIP (Supplementary Fig. S1B). These results support a model that tandem linkage of FIP and BIP is responsible for the false-positive LAMP reactions (Supplementary Fig. S1C).

### An improved bioinformatics algorithm for LAMP assay design

Based on the above experimental data for assay optimization, we developed an improved bioinformatics algorithm for designing LAMP primers for SARS-CoV-2 diagnosis. This algorithm incorporated many design features that were proven to be important for DNA amplification in our previous study ^21-23^. In addition, the new algorithm also included novel features that are unique for LAMP assays. As summarized in Fig. 3, we first downloaded all 106,439 SARS-CoV-2 genome sequences submitted to the GISAID database (https://www.gisaid.org) as of September, 2020. These genome sequences were aligned to identify consensus viral genome sequence. In our assays, we excluded target regions that contain any variable nucleotide (<98% consensus).

**Figure 3.**
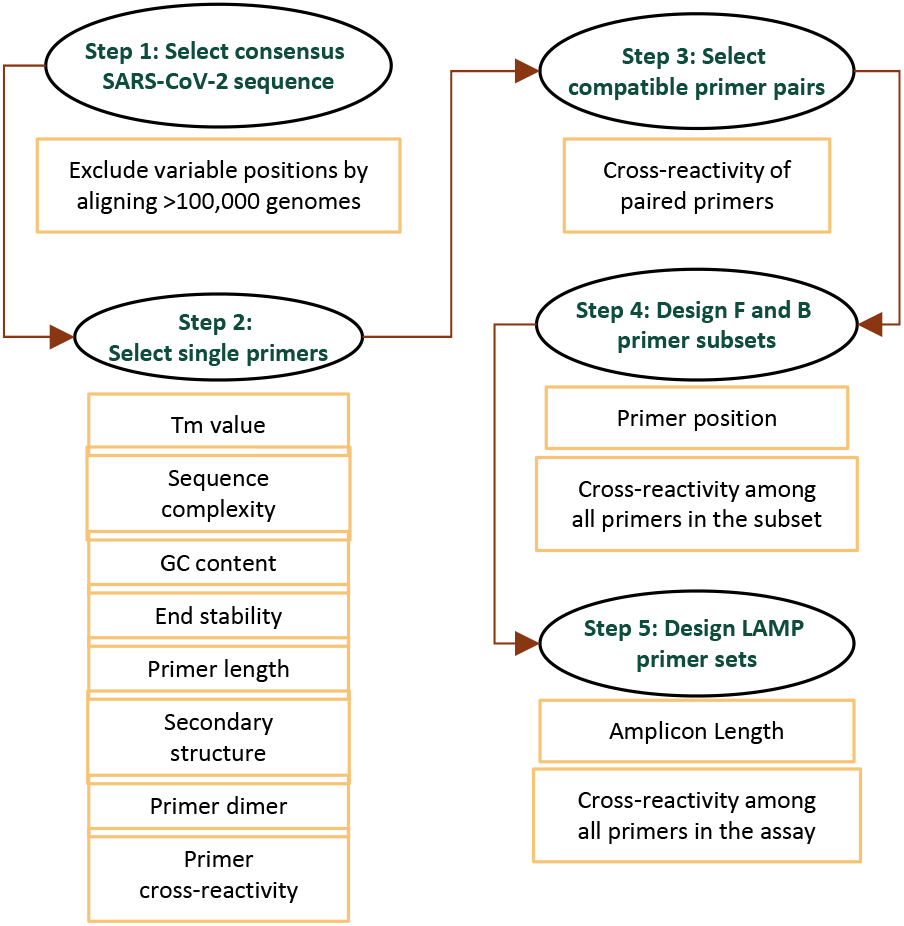
Workflow of the LAMP primer design algorithm.

Next, single primer candidates were selected from the target region, and a primer candidate would be discarded if any one of the following criteria was not met. The Tm value of the primer was in the range of 60-64°C, except for F1c/B1c candidates (Tm >45°C). All Tm values were calculated using the Nearest Neighbor method ^24, 25^. To avoid potential primer cross-reactivity due to low sequence complexity, a stretch of continuous same nucleotides was not allowed (4 C’s, 4 G’s, 5 A’s, or 5 T’s). To further exclude sequences of low complexity, the DUST program ^26^ was employed, and any sequence identified by DUST was rejected. In addition, the GC content of the primer was in the range of 35-65% to ensure uniform priming. The 3’-end of the primer contributes most to non-specific primer extension, especially if the binding of these nucleotides is relatively stable ^27^. Thus, the ΔG value of five residues at the 3’-end was calculated, and a threshold value of -8 kcal/mol was used for sequence rejection. The design algorithm further assessed potential secondary structures, which could hinder primer annealing to the template, leading to reduced amplification efficiency; primer secondary structures could also result in non-specific LAMP products by initiating unintended mispriming events. In our design, we implemented multiple secondary structure filters for primer screening as we described previously ^21-23^. In addition, the Mfold program ^28^ was employed to exclude primer candidates with a low ΔG value of predicted secondary structure.

After single primer candidates were selected, the design algorithm then chose compatible primer pairs that show no cross-reactivity based on sequence match ^21^. In particular, more stringent selection filters were applied to the end residues of paired primers to prevent the formation of primer heterodimers. Based on compatible primer pairs, the F and B primer subsets were selected, which were further assembled into a six-primer set for the LAMP assay. F1c and F2 were combined into the forward annealing primer (FIP); similarly, B1c and B2 were combined into the reverse annealing primer (BIP). Unintended primer cross-reactivity is a major challenge for DNA amplification, especially when multiple primers are included in the same reaction. To alleviate this concern, our algorithm includes filters to evaluate potential primer dimer formation or other mispriming events from all possible primer pairs among the six primers in the assay. Details of these cross-reaction filters were described previously ^21-23^. Moreover, restraints were placed on the relative positions of each primer in the assay for efficient LAMP amplification (Supplementary Fig. S2).

### Experimental validation of the RT-LAMP assays for SARS-CoV-2 RNA detection

The algorithm described above was used to design LAMP primers for SARS-CoV-2 diagnosis. As summarized in Fig. 4 and Supplementary Table S1, we tested the performance of 22 newly designed primer sets (N7-N28), as evaluated by both the TT value and FP score, to target the N gene sequence in a DNA plasmid. Most of these assays (n=19) had low FP scores (<=2), indicating high specificity of the LAMP reaction. Moreover, the majority of the assays (n=14) were able to detect the target with high efficiency, as demonstrated by low TT values (<30 min). In particular, N7, N25, and N27 assays had the best overall performance, with both TT value less than 20 min and FP score of 1. Thus, these three assays were further evaluated for their diagnostic potential using the SARS-CoV-2 RNA genome as the target.

**Figure 4.**
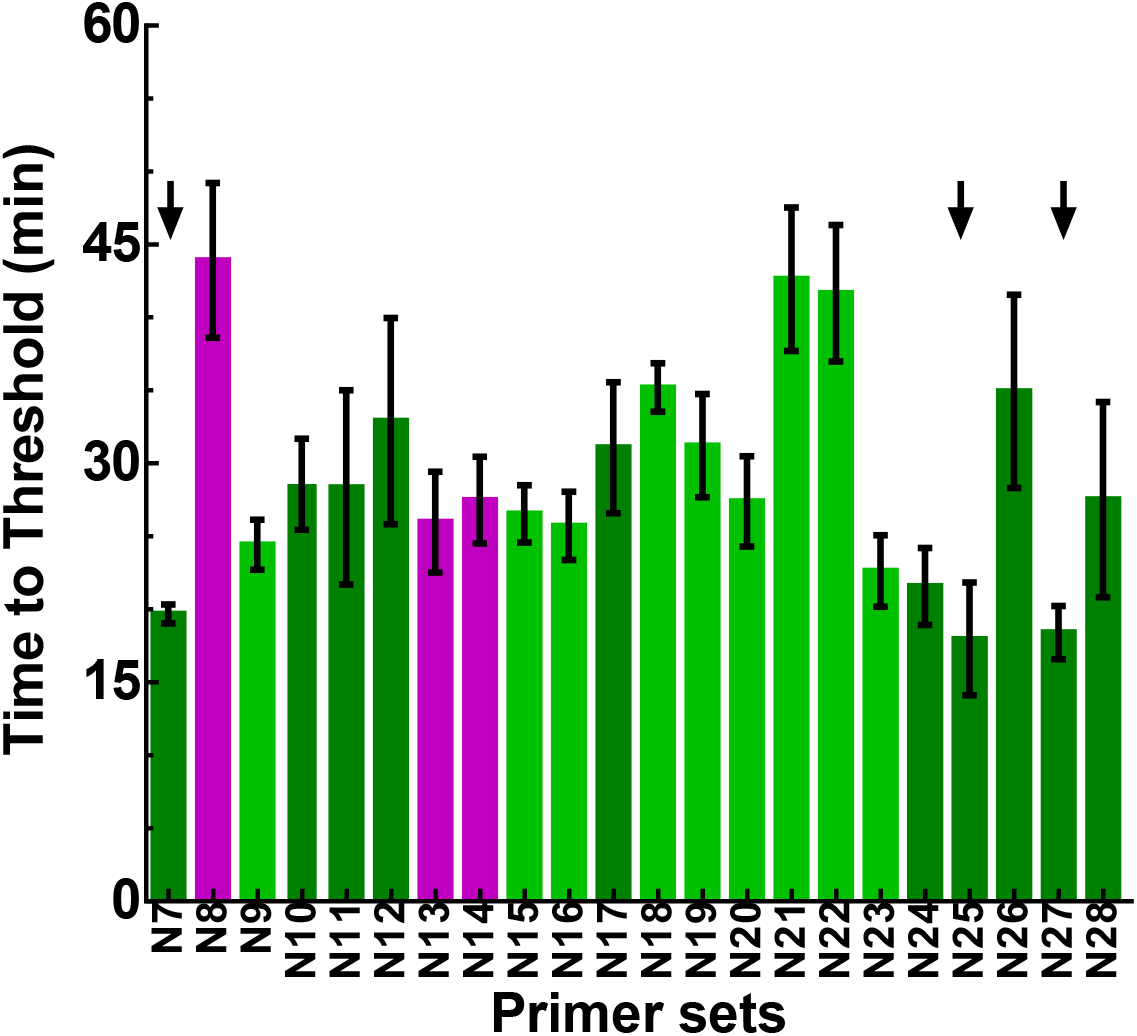
LAMP validation of designed primer sets targeting the N gene of SARS-CoV-2. The plasmid containing the N gene of SARS-CoV-2 (2,000 cp/reaction) was used as template for the LAMP reactions. The same color scheme as descried in Fig. 2B was used to represent the FP score. The arrows indicated selected LAMP assays with best overall performance in efficacy and specificity.

Besides primer sets for the N gene, we also designed and further tested six primer sets for the M gene and seven primer sets for the S gene, respectively, using SARS-CoV-2 RNA as the target (Fig. 5 and Supplementary Tables S3, S4 & S5). All the M gene assays were highly efficient at detecting SARS-CoV-2 RNA (TT value close to or below 30 min). Among them, M5 and M6 assays had the best overall performance (TT value close to or below 15 min with FP score of 1). Thus, these two assays were selected for further evaluation. As for the S gene assays, they had similar efficiency as compared to the M gene assays, but with elevated levels of non-specific amplification. Among them, S1, S4, and S7 assays had no observed non-specific products, with an FP score of 1. In the same way, we also tested selected N gene assays by RT-LAMP for viral RNA detection. Altogether, we identified five primer sets, N2(−5), N7, N27, M5, and M6, which had the best overall performance for both amplification efficiency and specificity (Fig. 5).

**Figure 5.**
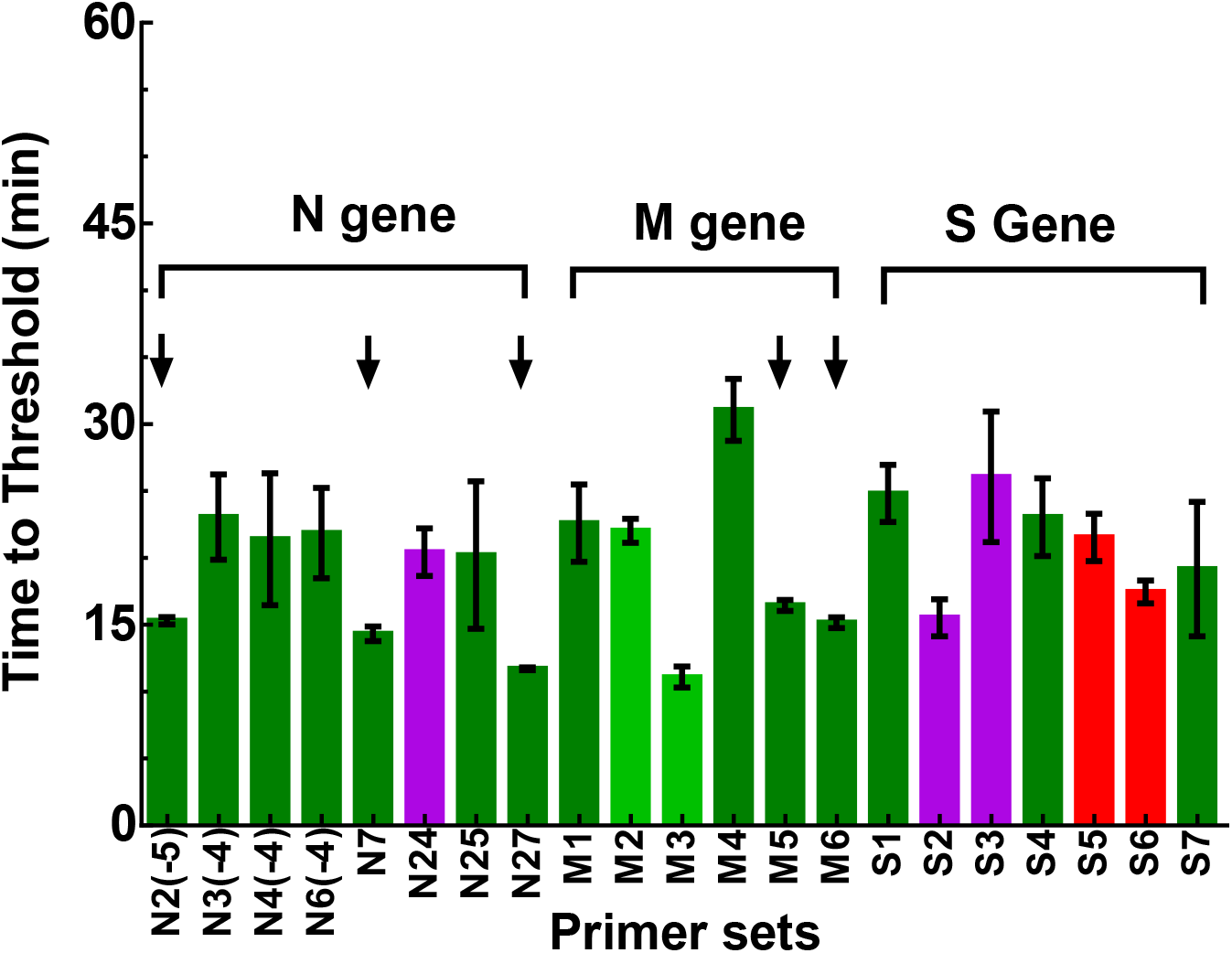
RT-LAMP validation of designed primer sets targeting the N, M, and S genes of SARS-CoV-2. SARS-CoV-2 RNA (2000 cp/reaction) was used as template for the RT-LAMP reactions. The same color scheme as descried in Fig. 2B was used to represent the FP score. The arrows indicated selected RT-LAMP assays with best overall performance in efficacy and specificity.

We further tested the detection sensitivity of five selected assays by serial dilution of SARS-CoV-2 RNA. The RT-LAMP results (Fig. 6A) showed a dose-dependent increase in TT values (from 11 to 65 min) with decreasing amounts of viral RNA in the reactions (from 10,000 to 1 cp/µL). All five assays could reliably detect 100 cp/µL of viral RNA in about 20 min or less (100% positivity). Interestingly, when the RNA amount was further diluted down to 20 cp/µL, N7, N27, and M6 assays maintained a 100% positive detection rate with the TT of 19-36 min. Further, when 5 cp/µL of RNA was tested, N7 assay had a positive rate of 80% (average TT of 43 min); N27 had a rate of 40% (average TT of 19 min); M5 had a rate of 55% (average TT of 49 min); and M6 had a rate of 53% (average TT of 41 min). When only 1 cp/µL of RNA was added to the reaction (based on serial dilution), the positive rate of viral detection was much lower for all these assays, ranging from 0-20%.

**Figure 6.**
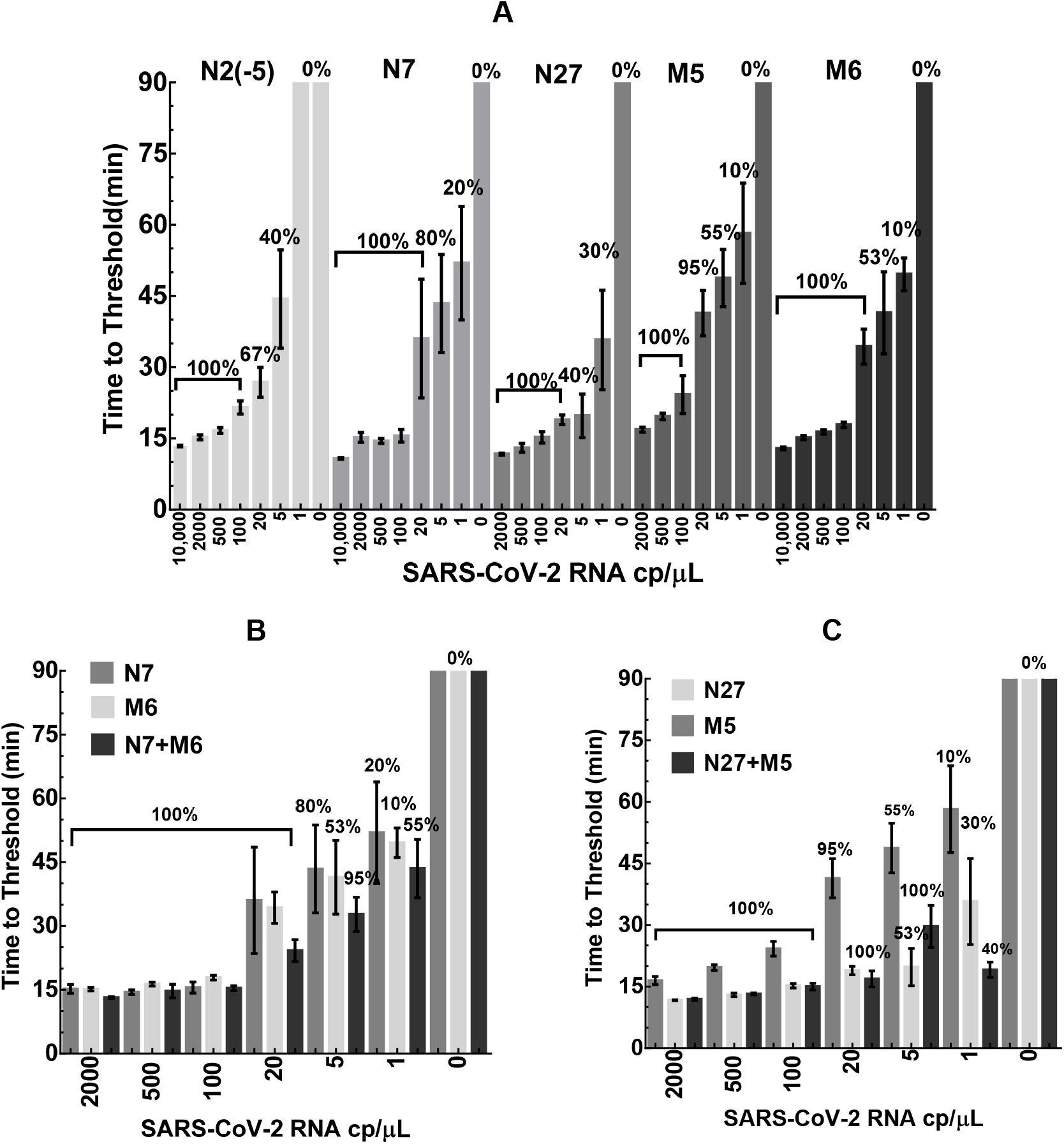
Sensitivity of the RT-LAMP assays in detecting SARS-CoV-2 RNA. SARS-CoV-2 RNA was diluted in water and added to the reactions as template (0-1,000 cp/µL). The TT values were determined by averaging 10-20 independent measurements and presented as mean±SD. The percentage of positive reactions among all replicated reactions for each assay was also presented. **(A)** single primer sets N2(−5), N7, and N27 for the N gene, as well as M5 and M6 for the M gene. **(B)** Multiplexed N7+M6 assay vs. N7 or M6 assay alone. **(C)** Multiplexed N27+M5 assay vs. N27 or M5 assay alone.

### Multiplexed RT-LAMP to further improve assay sensitivity

We next sought to determine whether RT-LAMP sensitivity could be further improved when assays were combined for simultaneous detection of multiple genes. Among the validated assays, N7/N27 and M5/M6 assays were designed to target the N gene and M gene, respectively. Thus, we established two multiplexed assays based on N7+M6 and N27+M5, respectively. Both assays had no detectable background (FP score of 1). Notably, as shown in Fig. 6B&C, the efficiency and sensitivity of the multiplexed assays (as evaluated by TT and detection rate) were significantly improved over any single assay alone. Specifically, N7/M6 assay could reliably detect 5 cp/µL of viral RNA in about 33 min (with a detection rate of 95% vs. 80% for N7 and 53% for M6, Fig. 6B). Similarly, N27/M5 assay could detect 5 cp/µL of RNA in 30 min (detection rate of 100% vs. 55% for M5 and 53% for N27, Fig. 6C). Most noticeably, with 1 cp/µL of RNA (based on serial dilution), both multiplexed assays had significantly improved detection rate compared with the single assays (40-55% detection rates for the multiplexed assays vs. 10-30% for the single assays).

For clinical diagnosis, the human samples (e.g., saliva) to be tested contain transcripts of both virus and human origin. To evaluate potential assay cross-reaction to human RNA, we spiked in various amounts of SARS-CoV-2 genomic RNA into total RNA (10 ng/µL) from HeLa cells or pretreated human saliva. Consistent with the data presented in Fig. 6B&C, the RT-LAMP results showed that multiplexed N7/M6 and N27/M5 assays were efficient and sensitive at detecting as few as 5 cp/µL of viral RNA (100% detection rate with TT <30 min, Fig. 7A&B) mixed with HeLa RNA. When only 1 cp/µL of viral RNA was tested, the detection rates of the multiplexed assays ranged from 70%-80% with the TT around 30 min. For pretreated saliva samples spiked with SARS-CoV-2 RNA, both assays showed similar but slightly decreased performance in sensitivity and efficiency compared with HeLa RNA (Fig. 7A&B). Importantly, the reactions produced negative results (i.e., 0% detection rate) when viral RNA was omitted, indicating no cross-reactivity of the assays to HeLa RNA or saliva RNA. As a positive control, we also performed RT-LAMP to target Beta-actin (ACTB) RNA in HeLa cells and human saliva (Supplementary Table S7)^29^, and the result indicated high efficacy of detection (TT ∼10 min for HeLa RNA and ∼16min for saliva RNA, respectively). We further evaluated colorimetric changes of the reactions for detecting target transcripts (Supplementary Fig. S3), which showed consistent results to the fluorescent readouts (Supplementary Fig. S3A & B).

**Figure 7.**
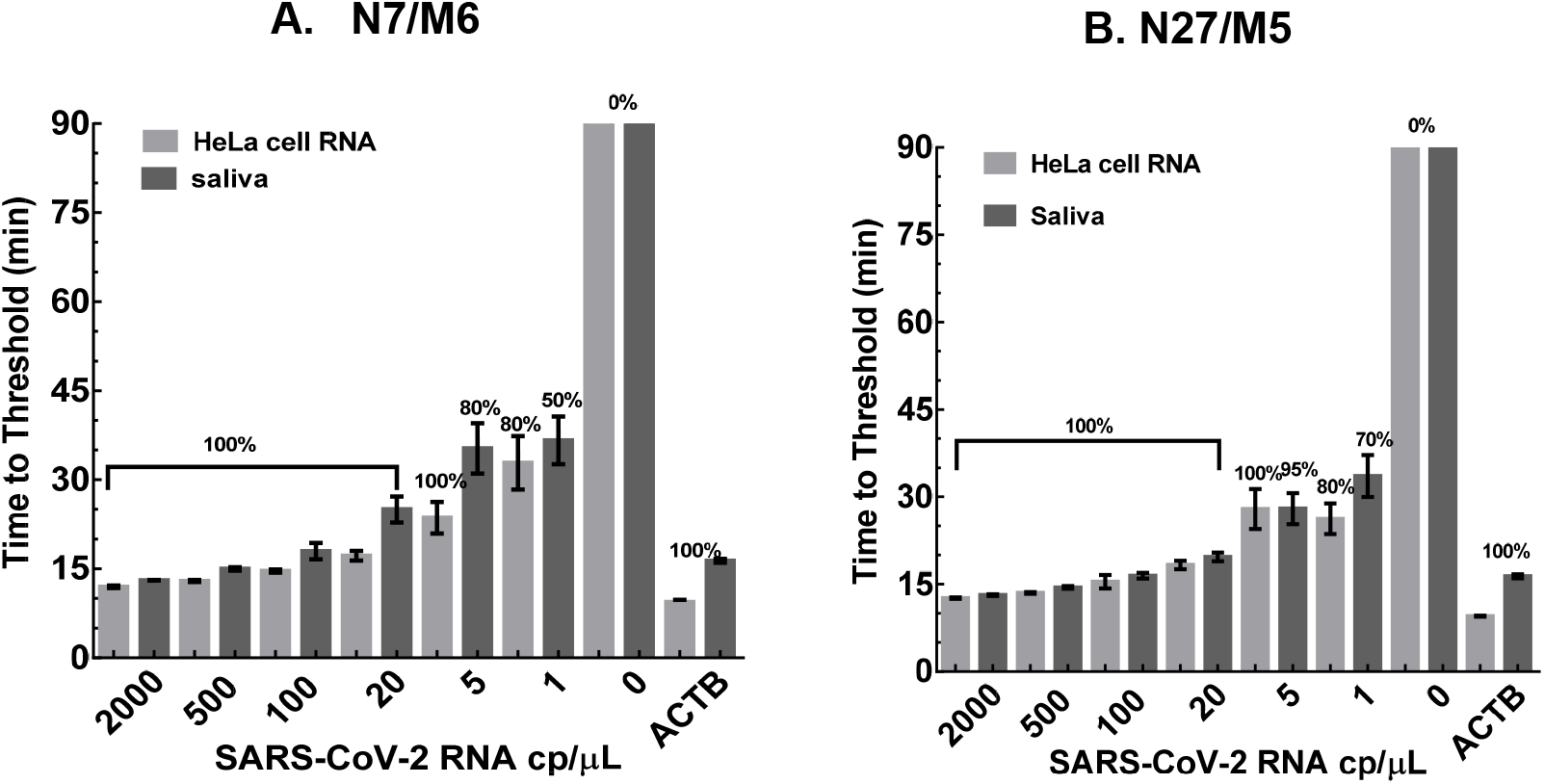
The sensitivity and specificity of two multiplexed assays in detecting SARS-CoV-2 RNA mixed with HeLa RNA or human saliva. SARS-CoV-2 RNA (0-1,000 cp/µL) was mixed with HeLa cell RNA (10 ng/µL) or pretreated healthy human saliva. The TT values were determined by averaging 10-30 independent measurements and presented as mean±SD. Beta-actin (ACTB) RT-LAMP assay was used as positive control for human RNA detection. The numbers of replicates were: 100-2,000 cp/µL reactions, n=5-10; 0-20 cp/µL reactions, n=20-30. **(A)** Validation of the N7/M6 assay. **(B)** Validation of the N27/M5 assay.

### Further optimization of the RT-LAMP assays for direct detection of SARS-CoV-2 in saliva

With the limit of detection (LOD, defined as ≥95% detection rate) at 5 cp/µL of viral RNA (Tables 2), N7/M6 and N27/M5 assays were highly efficient and specific when SARS-CoV-2 RNA was diluted with water or HeLa RNA. However, the LODs were relatively low for SARS-CoV-2 detection in saliva samples mainly due to the limited amount (0.5 µL) of pretreated saliva being added to the RT-LAMP mixture (the LODs at 800 cp/µL and 200 cp/µL of viral RNA in undiluted saliva for N7/M6 and N27/M5, respectively; Tables 2). To further increase the sensitivity for saliva testing, we increased the RT-LAMP reaction volume from 10 µL to 20 µL, such that more saliva could be added to the reaction. In addition, the amount of pretreated saliva being added to the reaction was optimized to 3 µL, according to a recent publication ^30^ as well as our testing data. Moreover, we observed that HPLC purification of the FIP and BIP primers further boosted the efficiency of RT-LAMP (data not shown).

**Table 2.** Limit of Detection (LOD) analysis of the RT-LAMP assays.

| **SARS-CoV-2 RNA with N7/M6 assay** |  |  |  |  |  |  |  |
| --- | --- | --- | --- | --- | --- | --- | --- |
| **Diluted in water** |  |  | **Mixed with HeLa RNA** |  | **Mixed with pretreated saliva** |  | **cp/μL in undiluted saliva** |
| **RNA cp/μL** | **Positive/ Replicates** | **Sensitivity** | **Positive/ Replicates** | **Sensitivity** | **Positive/ Replicates** | **Sensitivity** |  |
| 2000 | 10/10 | 100% | 10/10 | 100% | 5/5 | 100% | 80,000 |
| 500 | 10/10 | 100% | 10/10 | 100% | 5/5 | 100% | 20,000 |
| 100 | 10/10 | 100% | 10/10 | 100% | 5/5 | 100% | 4,000 |
| 20 | 20/20 | 100% | 20/20 | 100% | 30/30 | 100% | 800 |
| 5 | 19/20 | 95% | 20/20 | 100% | 24/30 | 80% | 200 |
| 1 | 11/20 | 55% | 16/20 | 80% | 15/30 | 50% | 40 |
| 0 | 0/20 | - | 0/20 | - | 0/30 | - | 0 |

| **SARS-CoV-2 RNA with N27/M5 assay** |  |  |  |  |  |  |  |
| --- | --- | --- | --- | --- | --- | --- | --- |
| **Diluted in water** |  |  | **Mixed with HeLa RNA** |  | **Mixed with pretreated saliva** |  | **cp/μL in undiluted saliva** |
| **RNA cp/μL** | **Positive/ Replicates** | **Sensitivity** | **Positive/ Replicates** | **Sensitivity** | **Positive/ Replicates** | **Sensitivity** |  |
| 2000 | 10/10 | 100% | 10/10 | 100% | 10/10 | 100% | 80,000 |
| 500 | 10/10 | 100% | 10/10 | 100% | 10/10 | 100% | 20,000 |
| 100 | 10/10 | 100% | 10/10 | 100% | 10/10 | 100% | 4,000 |
| 20 | 20/20 | 100% | 20/20 | 100% | 20/20 | 100% | 800 |
| 5 | 20/20 | 100% | 20/20 | 100% | 19/20 | 95% | 200 |
| 1 | 8/20 | 40% | 16/20 | 80% | 14/20 | 70% | 40 |
| 0 | 0/20 | - | 0/20 | - | 0/20 | - | 0 |

| **Saliva spiked with SARS-CoV-2 particles** |  |  |  |  |  |
| --- | --- | --- | --- | --- | --- |
| **N7/M6 assay (20 μL reaction)** |  |  | **N27/M5 assay (20 μL reaction)** |  |  |
| **Virus /uL** | **Positive/ Replicates** | **Sensitivity** | **Positive/ Replicates** | **Sensitivity** | **cp/uL in undiluted saliva** |
| 5 | 19/20 | 95% | 20/20 | 100% | 67 |
| 2.5 | 14/20 | 70% | 16/20 | 80% | 33 |
| 1.2 | 8/20 | 40% | 12/20 | 60% | 16 |
| 0.6 | 6/20 | 30% | 6/20 | 30% | 8 |
| 0 | 0/20 | - | 0/20 | - | 0 |

Under these optimized assay conditions for saliva analysis, we tested the performance of the N7/M6 and N27/M5 assays by directly adding heat-inactivated SARS-CoV-2 particles into saliva (i.e., “simulated” COVID-19 samples). Both assays had an LOD of 67 cp/µL of viral particles in simulated saliva samples. Below the LOD threshold, the N7/M6 assay had a positivity rate of 70% for 33 cp/µL, 40% for 16 cp/µL, and 30% for 8 cp/µL of SARS-CoV-2 in undiluted saliva samples, respectively. Similarly, the N27/M5 assay had a positivity rate of 80% for 33 cp/µL, 60% for 16 cp/µL, and 30% for 8 cp/µL of virus, respectively (Tables 2). Besides the improvement in LOD, assay time was also shortened to 70 min since viral RNA was detected in all samples within this timeframe while all negative control samples (0 cp/µL) remained negative for viral detection (data not shown).

### Performance comparison of the RT-LAMP assays using simulated and patient saliva samples

To evaluate the clinical utility of the RT-LAMP assays, we compared three alternative methods for SARS-CoV-2 diagnosis with saliva samples. **1)** RT-LAMP directly using saliva; **2)** RT-LAMP using RNA isolated from saliva; and **3)** RT-qPCR using RNA isolated from saliva. Currently, RT-qPCR is the most widely used method in the clinic for SARS-CoV-2 diagnosis mainly due to its high detection sensitivity and specificity. We evaluated a commonly used RT-qPCR assay, E-Sarbeco, which is recommended for COVID-19 diagnosis by the WHO and CDC ^31^. Our validation data indicated that the E-Sarbeco assay had an LOD of 0.5 cp/µL (i.e., 14 cp/reaction) of SARS-CoV-2 RNA, while producing no false positive signals from the negative control (Ct >40). This result is consistent with previously reported data on the E-Sarbeco assay and is also in the same LOD range with other RT-qPCR assays for COVID-19 diagnosis ^3, 32, 33^.

The experimental design for assay comparison was summarized in Fig. 8A. Both N7/M6 and N27/M5 assays reliably detected SARS-CoV-2 at 78 cp/µL or higher in simulated saliva samples (4 out of 4 repeats), using either saliva or RNA isolated from the saliva (Fig. 8B). With 5-39 cp/µL viral particles added to the saliva, both RT-LAMP assays consistently produced positive results (1-3 positive reactions out of four replicates) when tested directly on the saliva. In comparison, the sensitivity of the assays using isolated saliva RNA was significantly higher, as almost all reactions were positive at the same viral concentrations (Fig. 8B). At 1 cp/µL level, N7/M6 and N27/M5 assays produced 0 and 1 positive reaction (out of four replicates), respectively. In contrast, the same two assays produced two and three positive reactions, respectively, when tested on isolated saliva RNA. As reference control, we also performed RT-qPCR using the E-Sarbeco assay, and observed positive results across all viral concentrations, with Ct values in the range of 29.3 to 36.4 (Fig. 8B).

**Figure 8.**
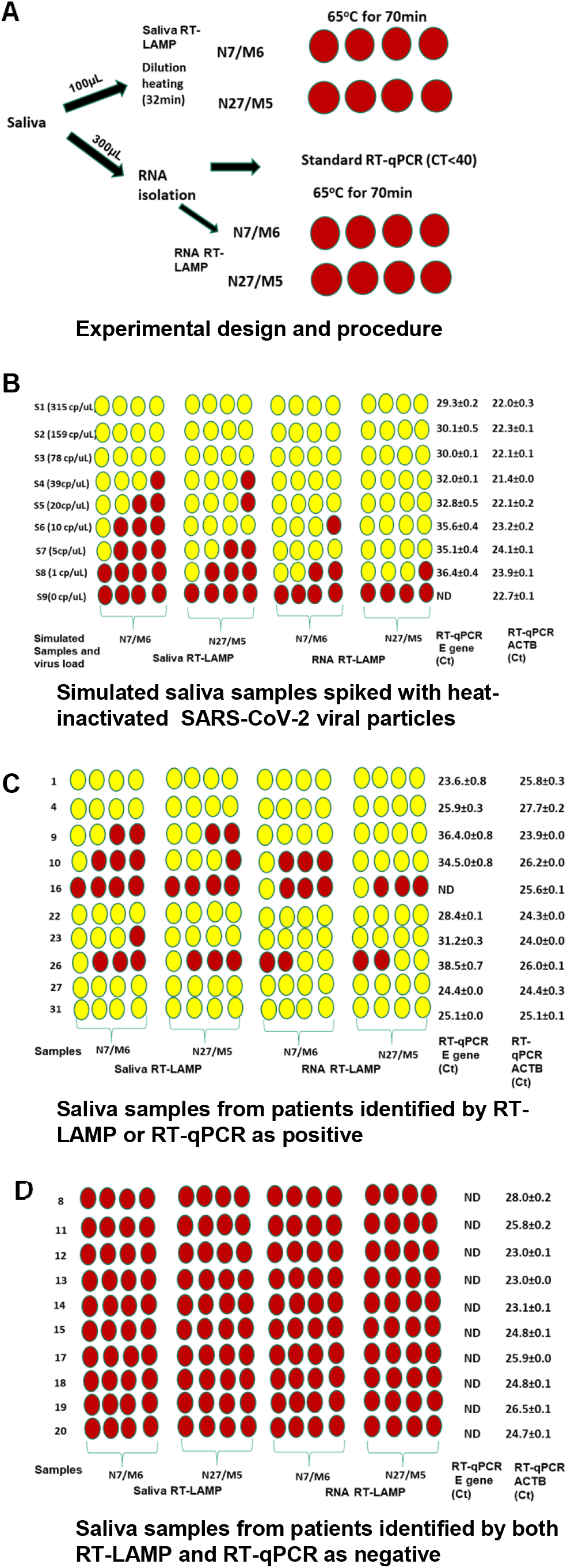
Validation of the RT-LAMP assays using simulated or clinical saliva samples containing SARS-CoV-2 virus. RNA isolation, saliva treatment, RT-LAMP, and RT-qPCR were performed as described in Methods. RT-LAMP was performed on either isolated saliva RNA or directly on heat-inactivated saliva. An RT-qPCR assay was also included for performance comparison. Schematic colorimetric results were presented, with yellow and red representing positive and negative reactions, respectively. **(A)** Overall study design and experimental procedure. **(B)** Simulated saliva samples spiked with SARS-CoV-2 particles at various concentrations (0-315 cp/µL). **(C)** Saliva samples from the patients identified by RT-LAMP or RT-qPCR as positive for SARS-CoV-2. **(D)** Saliva samples from the patients identified by both RT-LAMP and RT-qPCR as negative for SARS-CoV-2.

We further validated the two RT-LAMP assays using twenty clinical saliva samples originally acquired for COVID-19 diagnosis. Similar to the testing procedure for simulated saliva samples, we evaluated the performance of the RT-LAMP assays using both saliva and RNA isolated from saliva. As reference control, the E-Sarbeco RT-qPCR assay was also included for testing of saliva RNA. Out of the twenty cases, nine were tested positive (Fig. 8C, patients #1, 4, 9, 10, 22, 23, 26, 27, and 31), and ten were tested negative (Fig. 8D, patients #8, 11, 12,13, 14, 15, 17, 18, 19, and 20) by all three testing methods. The viral concentrations in these positive samples varied greatly, as indicated by a broad range of Ct values (23.6-38.5) from RT-qPCR. Interestingly, one patient (#16) was tested negative by RT-qPCR and also by saliva RT-LAMP (i.e., direct test on saliva). However, this patient was tested positive by both N7/M6 and N27/M5 assays using isolated saliva RNA (one positive reaction out of four replicates). High-throughput sequencing of the resultant RT-LAMP products confirmed specific amplicon sequences from SARS-CoV-2. (Supplementary Table S6).

## Discussion

RT-LAMP is an emerging technology for molecular diagnosis of human pathogens. Compared with RT-qPCR, the advantages of RT-LAMP assays are clear: 1) The reaction is fast; 2) the assay setup does not require any expensive instrument; and 3) the reaction is relatively insensitive to sample impurity, making it possible to directly test crude samples without RNA isolation. Thus, RT-LAMP with colorimetric readout is particularly attractive for point-of-care diagnosis of infectious pathogens. For example, multiple investigators have attempted to develop RT-LAMP assays for the diagnosis of COVID-19 ^30, 34^.

Despite its potential as a powerful diagnostic tool, RT-LAMP has not been widely adopted in the clinical setting (in comparison of RT-qPCR). One major challenge is the design of robust LAMP primers. A LAMP assay includes a set of six primers, targeting eight regions of the nucleic acid sequence for amplification; thus, the primer design is complicated. Existing assay design methods often produce ineffective primer sets due to poor amplification efficiency or non-specific amplification. As a result, high-quality LAMP assays are commonly developed by trial and error based on experimental testing of many designed primer sets. Due to the major challenges in LAMP primer design, most LAMP studies for COVID-19 diagnosis adopted previously published primer sets that had suboptimal performance ^11, 30^.

To address the aforementioned challenges, we developed an improved algorithm for LAMP primer design to target SARS-CoV-2. First, we optimized the length of F1c/B1c to significantly boost the amplification efficiency. F1c and B1c make up the loop stem of the dumbbell structure, which is formed at the beginning of the LAMP reaction and serves as template to initiate the amplification phase (Fig. 1). Second, Next-generation sequencing demonstrated that false-positive reactions mainly resulted from unintended interactions between FIP and BIP, two longest primers that are required at much higher concentrations than other primers in the reaction. Thus, more stringent specificity filters should be applied to the design of FIP/BIP primers. Taking account of these LAMP-specific parameters as well as other common primer design criteria from our previous studies ^21-23^, we developed an improved bioinformatics algorithm for LAMP primer design. Our design method is highly effective at producing high-quality LAMP assays, as evidenced by the validation data for the primer sets targeting the N and M genes of SARS-CoV-2. However, for the S gene, it is still a challenge to design efficient primer sets, likely due to the higher GC content (58% for S vs.43% for M and 47% for N).

Saliva testing is an attractive method for COVID-19 diagnosis. 1) It is a non-invasive method for sample collection; 2) saliva can be collected by a non-medical care worker, thus reducing the risk of nosocomial transmission; and 3) saliva is relatively stable at a broad range of storage temperatures ^35, 36^. Despite these advantages, there are also challenges facing COVID-19 saliva testing: 1) saliva has a broad range of virus loads (10-100,000 cp/µL) ^37^; 2) saliva contains many heterogeneous components, some of which are inhibitory to most diagnostic assays. As a result, RNA isolation is required for standard RT-qPCR methods for COVID-19 diagnosis with saliva samples.

The RT-LAMP assays we developed could directly detect the SARS-CoV-2 virus in saliva samples and were highly sensitive to cover a broad range of SARS-CoV-2 titers in patient saliva (10-100,000 cp/µL). Importantly, our RT-LAMP assays produced identical diagnostic results to RT-qPCR when directly applied to patient saliva samples. Interestingly, SARS-CoV-2 titers in these saliva samples varied greatly based on the RT-qPCR results. As for low virus titers, it is a challenge to sensitively detect the virus, whether using RT-LAMP or RT-qPCR. However, this is not likely a major issue for curbing the spread of COVID-19; multiple studies showed that, when the virus titer was below a certain level (<100-1,000 cp/µL in biofluid or Ct value >34), the patients are not likely to be spread disease ^38-41^.

Our RT-LAMP assays were fast as most reactions were completed within 30-45 min. Moreover, the assays could be easily adapted for point-of-care diagnosis with colorimetric readout under constant reaction temperature, or as a high-throughput screening assay in 384-well plate format. In term of recently emerged SARS-CoV-2 variants, most identified mutations are located in the S gene sequence ^42, 43^, and thus our validated assays focusing on the N and M genes will not likely be compromised by these mutations. In summary, we developed an improved algorithm for LAMP assay design, and further validated the assays targeting SARS-COV-2. The design algorithm as well as the validated RT-LAMP assays provide powerful diagnostic tools to curb the pandemic of COVID-19.

## Methods

### Assay targets for SARS-CoV-2 diagnosis

This COVID-19 research was approved by the Institutional Review Board at the University of Illinois at Chicago. The plasmid containing the N gene sequence of SARS-CoV-2 was purchased from Integrated DNA Technologies (IDT; 10006625, CoV-19 positive control). Synthetic SARS-CoV-2 genomic RNA [referred by the U.S. Food and Drug Administration (FDA)] was purchased from Twist Bioscience (102024, sequence based on GenBank accession MN908947.3). Heat-inactivated SARS-CoV-2 particles were acquired from the ATCC (VR-1986HK, 4.2×10^5^ particles/µL).

### LAMP and RT-LAMP assays

All DNA oligo primers were purchased from Sigma-Aldrich (desalt grade or as indicated). The N gene plasmid or SARS-CoV-2 RNA was diluted with DNase/RNase-free water or mixed with HeLa cell total RNA (10 ng/µL). The LAMP or RT-LAMP reactions were assembled in 384-well plate (Applied Biosystems). Each 10 µL reaction contained 5 µL of 2X WarmStart Colorimetric Master Mix (New England Biolabs or NEB, M1800), 0.5 µL of 20X SYBR Green I Nucleic Acid Gel Stain (5 µM, Life Technologies), 1 µL of primer set mix (final concentrations of 1.6 µM for FIP or BIP primer, 0.4 µM for FL or BL primer, 0.2 µM for F3 or B3 primer), 0.5 µL of guanidine hydrochloride (840 mM), 0.5 µL of target RNA or DNA, and 2.5 µL of DNase/RNase-free water. The LAMP or RT-LAMP assay in 10 µL reaction volume was used for primer screening and initial saliva assay optimization. A 20 µL reaction volume with the same reagent concentrations was subsequently adopted for assay validation using simulated and clinical saliva samples. For each 20 µL reaction, 10 µL of mineral oil was included to overlay the reaction mixture before initiating the reaction. When combining two LAMP primer sets in a single reaction, the concentration of each FIP or BIP primer was adjusted to 1.0 µM, while the concentrations of other primers remained the same. All LAMP and RT-LAMP reactions were performed in a real-time PCR instrument (Applied Biosystems QuantStudio 5) for 70-100 min at 65°C, with fluorescence signals acquired as fixed intervals. The colorimetric change of the reaction (i.e., positive samples turning yellow from pink) was recorded by a phone camera at the end of the reaction. The efficacy of LAMP or RT-LAMP amplification was evaluated by detection of fluorescence signals over a threshold readout (i.e., Time to Threshold, or TT). Optimal threshold was established using QuantStudio Design and Analysis Software 1.5.1 (Applied Biosystems). Readings from 5-10 replicated reactions or as indicated were averaged and the results were presented as mean±SD.

### RT-qPCR assays

E-Sarbeco, a widely used qPCR primer set targeting the E gene of SARS-CoV-2 ^11, 32^ and recommended by the CDC, was included in RT-qPCR reactions as standard diagnosis method for COVID-19 (Supplementary Table S8). RT reactions were performed with the High-Capacity cDNA Reverse Transcription Kit (Applied Biosystems). Each 10 µL of RT reaction included 1 µL of 10X RT buffer, 0.4 µL of 25X dNTP (100 mM), 1 µL of 10X random primers, 0.5 µL of reverse transcriptase, 1-5 µL of the RNA sample, and water to final volume of 10 µL. The RT reaction mixture was incubated at 25°C for 20 min followed by 37°C for 60 min, and finally heat inactivated at 85°C for 5 min. Real-time PCR was performed with Power SYBR Green Master Mix (Applied Biosystems). Each 14 µL reaction included 1 µL of diluted RT product, 7 µL of 2X Power SYBR Green Master Mix, 6 µL of the primer mix (final concentration at 250 nM for each E-Sarbeco primer). The real-time PCR running protocol was 10 min at 95°C, followed by 40 amplification cycles (95°C for 10 s and 60°C for 30 s).

### HeLa cell RNA

HeLa cells were cultured at 37°C with 5% CO_2_ in DMEM media supplemented with 10% FBS. Total RNA was isolated using the mirVana kit (Life Technologies) according to recommended protocol.

### Processing of saliva samples

Patient saliva samples were previously collected for COVID-19 diagnosis at a UIC clinical lab. In addition, we also analyzed pooled saliva samples from healthy individuals (Innovative Research). Saliva samples were first diluted with 0.9X volume of TE buffer (10 mM Tris, 1 mM EDTA pH 8.0). Then, 0.1X volume of 1X RNAsecure (25X, ThermoFisher) and 16 units/mL protease K (800 units/ml, NEB) were added to make 1:1 dilution of the saliva sample. The reaction mixture was incubated at 37°C for 10 min, 65°C for 10 min, and finally 95°C for 12 min to inactivate protease K. For spike-in experiments, heat-inactivated SAR-CoV-2 virus particles were added to the saliva before TE dilution. For patient saliva samples (n=20), they were first heated at 65°C for 30 min to inactivate the SARS-CoV-2 virus, and then further processed as described above. Four replicated reactions were performed directly for each saliva sample using two independent RT-LAMP assays. In addition, for each saliva sample, an aliquot (300 µL) was used for RNA isolation with the MagMAX Viral/Pathogen Nucleic Acid Isolation Kit (A42352, Applied Biosystems), followed by RT-qPCR or RT-LAMP analysis.

### Sequencing of LAMP products

DNA sequencing libraries were prepared by PCR amplification of LAMP or RT-LAMP products with modified universal primers for Illumina sequencing. The indexed primers contain the P5 or P7 sequence for Illumina sequencing at the 5’-end, a 9-bp index sequence in the middle, followed by an adaptor sequence and a 3’-end F2 or B2 sequence specific to the LAMP assay. The PCR reaction was set up by mixing 1 µL of diluted LAMP product, 1 µL of primer mix (10 µM), 12.5 µL of 2X PCR master mix (Tag Plus Master Mix Red, Lamda Biotech), and 10.5 µL of water. The PCR was performed at 94°C for 10 s, followed by 35 cycles of amplification (94°C for 10 s and 60°C for 45 s), and then 72°C for 1 min. The PCR products were purified with Ampure beads (Agencourt) and then quantified with QuantiFluor (Promega). The library samples were sequenced on MiniSeq (Illumina). The sequencing reads were aligned to the SARS-CoV-2 genome to identify virus-specific sequences. In addition, BLAST was performed to identify any sequence match to the NCBI GenBank database.

## Data Availability

Supplemental data are available at the journal website. The sequencing data are available at NCBI GEO under accession GSE173086.

## Data availability

Supplemental data are available at the journal’s website. The sequencing data are available at NCBI GEO under accession GSE173086.

## Author contributions

X.W. conceived and supervised the research. X.H. conducted the experiments. X.H. and G.T. performed the data analysis. N.I. helped on clinical validation of the methods. X.H., G.T., N.I., and X.W. interpreted the data and wrote the paper.

