## Supplementary figures for "Developing RT-LAMP Assays for Detection of SARS-CoV-2 in Saliva"

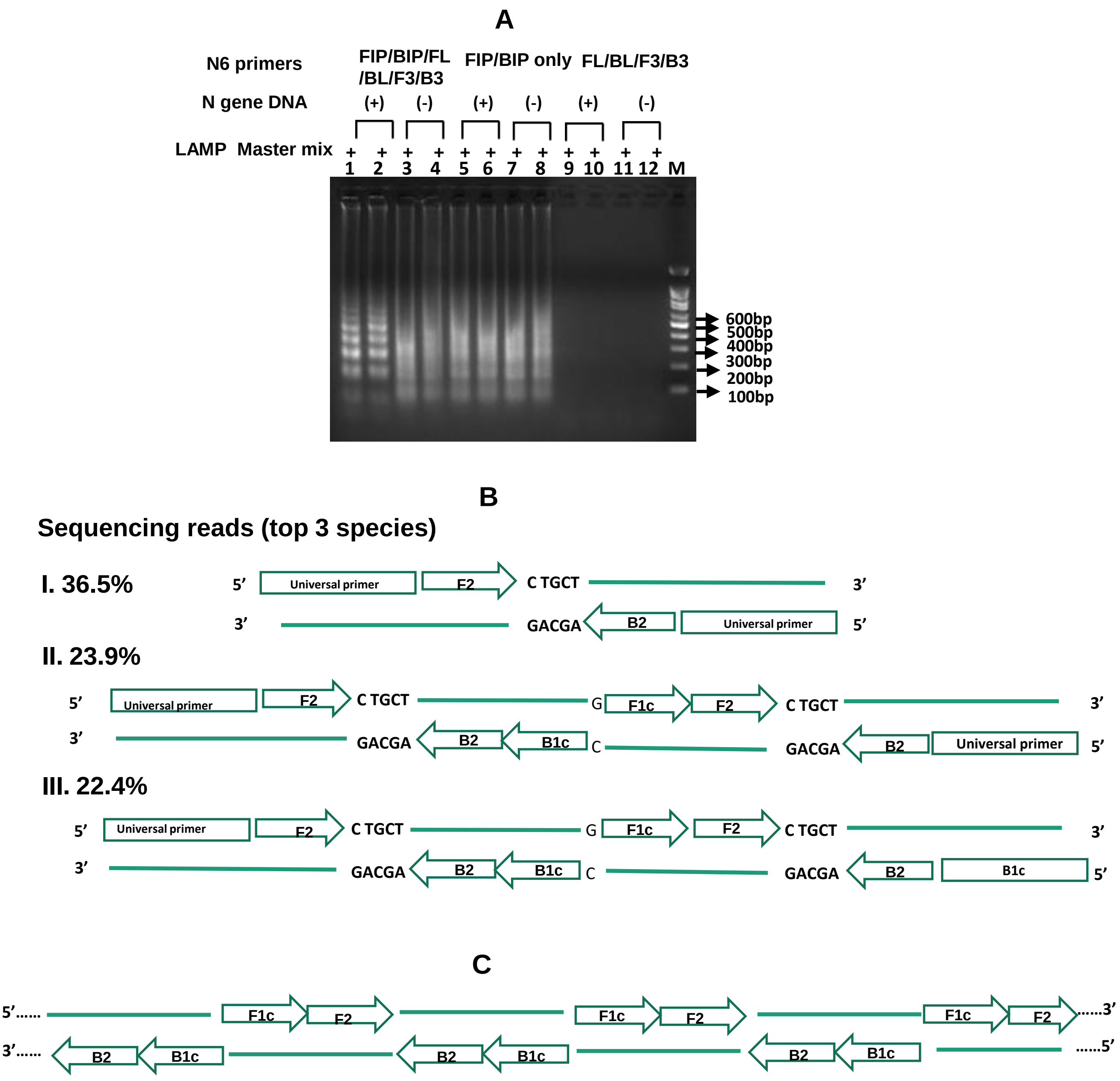


**Figure S1. Analysis of the false-positive products from the N6 assay. (A)** Gel electrophoresis of the LAMP products from the reactions, in the presence or absence of the template DNA (the plasmid containing the N gene sequence of SARS-CoV-2) (lanes 1-12): in the presence of full set primers (lanes 1-4); in the presence FIP/BIP primers only (lanes 5-8), or in the presence of other primers only (reactions 9-12). **(B)** Illumina sequencing analysis of the non-specific products as shown in lanes 3-8. **(C)** Hypothesized model for the formation of non-specific LAMP products.

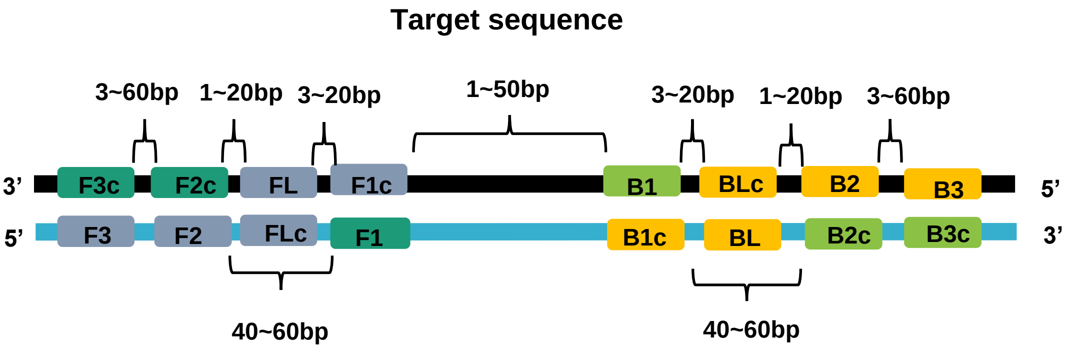


**Primer Length**

| bp | F3 | F2 | FL | F1 | B1 | BL | B2 | B3 | FIP(F2+F1c) | BIP(B2+B1c) |
|---|---|---|---|---|---|---|---|---|---|---|
| Min | 19 | 19 | 19 | 12 | 12 | 19 | 19 | 19 | | |
| Max | 25 | 25 | 25 | 17 | 17 | 25 | 25 | 25 | 37 | 37 |
| Opti | 22 | 22 | 22 | | | 22 | 22 | 22 | | |

**Figure S2. Restraints on the relative position and length of the LAMP primers.**

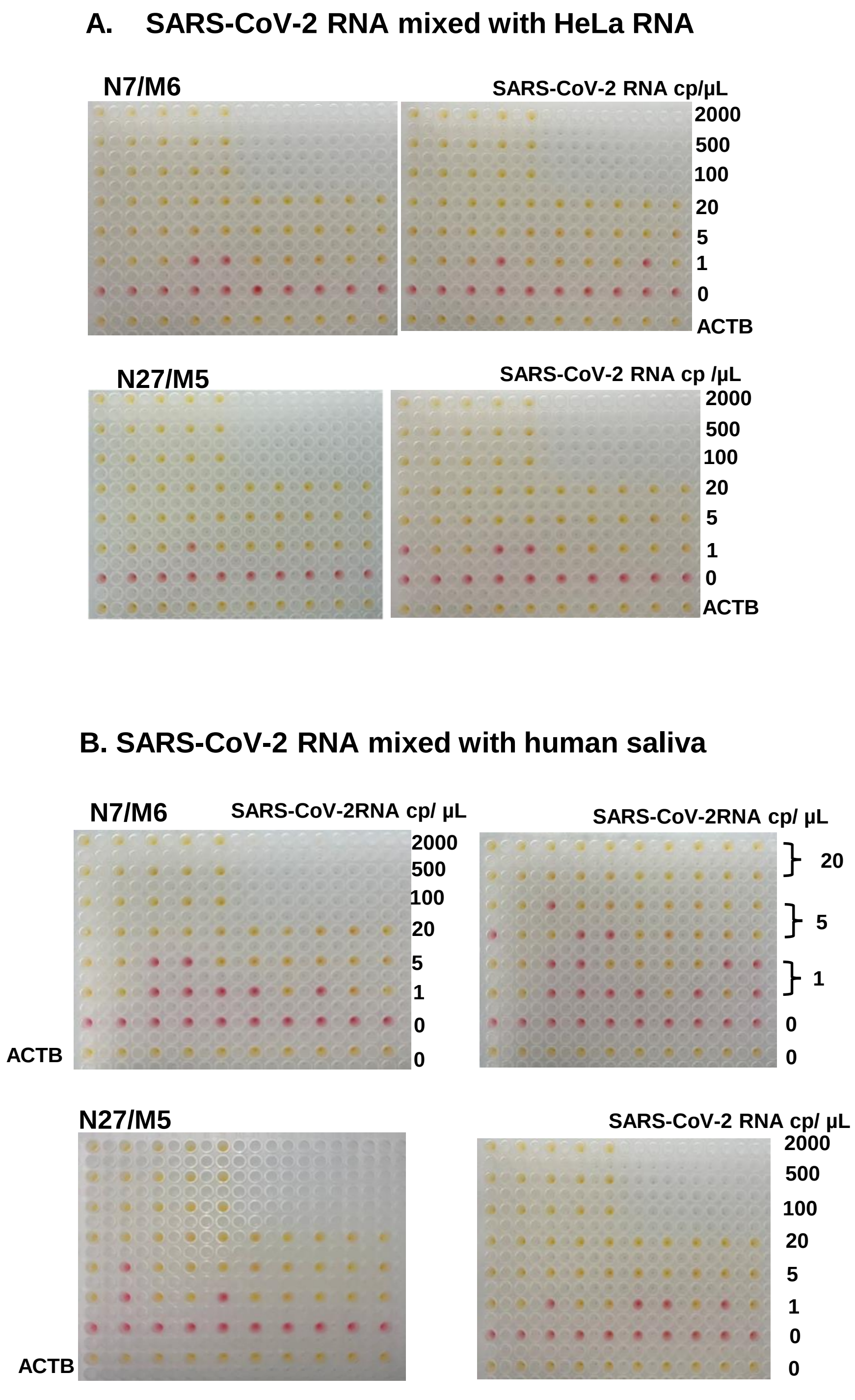


**Figure S3. Sensitivity and specificity of the RT-LAMP reactions assessed by colorimetric readout.** N7/M6 and N27/M5 assays were applied to various concentrations of SARS-CoV-2 RNA mixed with **(A)** HeLa cell RNA or **(B)** pretreated saliva from healthy humans. The concentrations of the SARS-CoV-2 RNA in the reaction were indicated. Yellow: positive; pink: negative; ACTB: Beta-actin control.
